# A Machine Learning Model Reveals Older Age and Delayed Hospitalization as Predictors of Mortality in Patients with COVID-19

**DOI:** 10.1101/2020.03.25.20043331

**Authors:** Jit Sarkar, Partha Chakrabarti

## Abstract

**Objective:** The recent pandemic of novel coronavirus disease 2019 (COVID-19) is increasingly causing severe acute respiratory syndrome (SARS) and significant mortality. We aim here to identify the risk factors associated with mortality of coronavirus infected persons using a supervised machine learning approach.

**Research Design and Methods:** Clinical data of 1085 cases of COVID-19 from 13^th^ January to 28^th^ February, 2020 was obtained from Kaggle, an online community of Data scientists. 430 cases were selected for the final analysis. Random Forest classification algorithm was implemented on the dataset to identify the important predictors and their effects on mortality.

**Results:** The Area under the ROC curve obtained during model validation on the test dataset was 0.97. Age was the most important variable in predicting mortality followed by the time gap between symptom onset and hospitalization.

**Conclusions:** Patients aged beyond 62 years are at higher risk of fatality whereas hospitalization within 2 days of the onset of symptoms could reduce mortality in COVID-19 patients.

## INTRODUCTION

The recent pandemic of coronavirus disease 2019 (COVID-19), caused by the severe acute respiratory syndrome coronavirus 2 (SARS-CoV-2) has caused unprecedented morbidity and mortality in almost all the continents (1). Despite implementations of extensive control measures, spread of the disease and eventual fatality could not be effectively halted till date. The major cause of death in COVID-19 is due to virus-induced pneumonia leading to respiratory failure (2). Epidemiological evidence suggests that older age and the associated co-morbidities such as cardiovascular disease and diabetes put patients at higher risk of mortality (3). Thus identification of novel risk factors predictive for patients’ outcome including mortality is needed.

Here using the publicly available clinical data from Kaggle, we have employed a machine learning tool to identify the risk factors that could potentially contribute to the mortality of COVID-19 patients from 22 countries in 4 continents. We show that older age and delayed hospitalisation of symptomatic patients are the two major risk factors for mortality in COVID-19 patients.

## MATERIALS AND METHODS

### Data source and preparation of dataset

The dataset was downloaded from Kaggle (https://www.kaggle.com/sudalairajkumar/novel-corona-virus-2019-dataset#COVID19_line_list_data.csv) on 23^rd^ March, 2020. It contained a total of 1085 reported cases of COVID-19 from 13^th^ January to 28^th^ February, 2020. Missing values were removed for all the variables to obtain a dataset of 433 individuals. 3 cases were filtered out from the dataset as the date of hospital visit preceded the date of symptom onset for them. Among the 430 cases selected finally from 22 countries in Asia, Australia, Europe and North America, there were cases of 37 deaths and 78 recoveries. The descriptive statistics of the deaths and confirmed recovered cases have been depicted in Table 1.

**Table 1.**
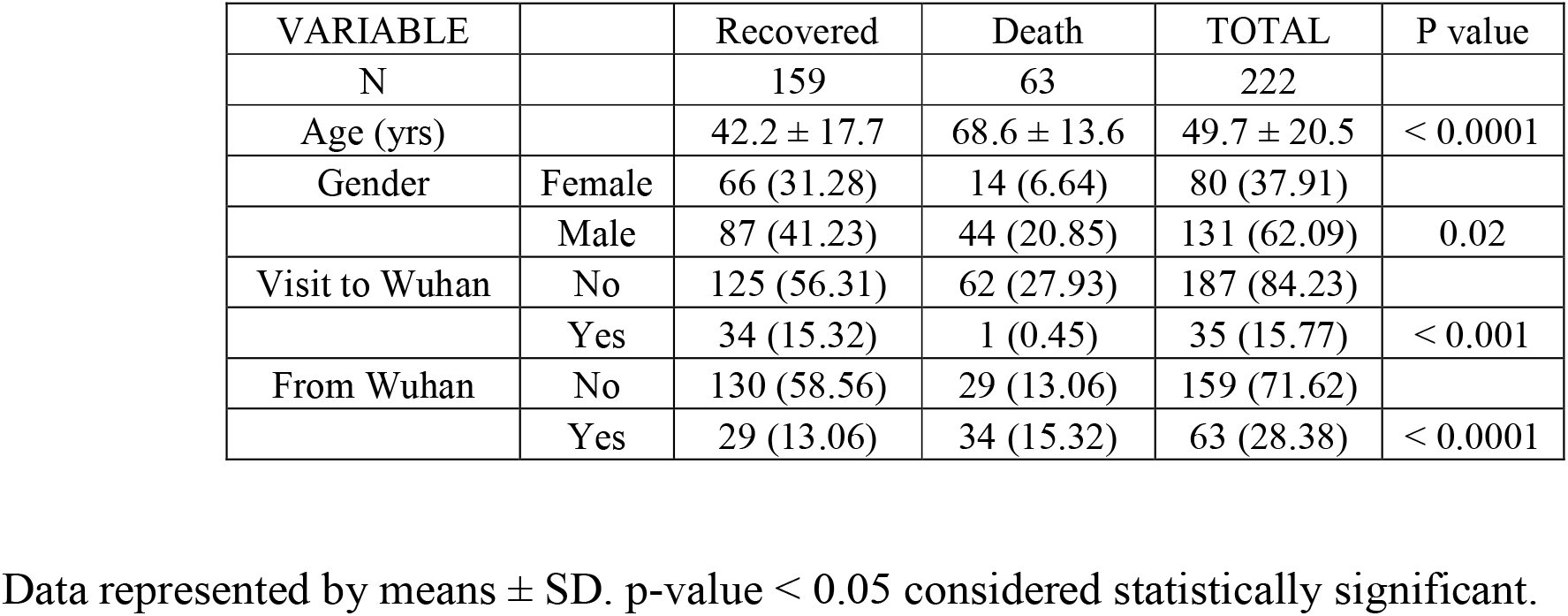
The subject characteristics of COVID-19 patients with Death and confirmed Recovery.

### Machine Learning Algorithm and Statistical Analysis

Random Forest classification algorithm (4) was implemented over a dataset with 37 deaths and 78 recoveries using the *randomForest* package in R. The dataset was randomly split into training and test dataset containing 70% and 30% of the total samples respectively. To evaluate the model performance, the Area under the ROC curve was calculated on the test dataset. A variable importance plot was generated using the importance of the predictors over the outcome. The importance of the variables has been reported according to both the mean decrease of Gini and the mean decrease of Accuracy. The partial dependency plots were finally generated using the *pdp* package in R to determine the marginal effect of the Age and Time to Hospitalization over the fate of COVID-19 infection.

The descriptive summary of the data has been represented by mean and standard deviation (SD). The numerical variables have been compared between groups by independent-samples two-sided Student’s t-test. The categorical variables have been tested using Chi-square test. All the statistical analyses were performed in RStudio (Version 1.2.1335) (5).

## RESULTS & DISCUSSIONS

Out of total 1085 reported cases of COVID-19 infection, the Random Forest model was built over the 70% of the dataset of 115 patients (37 deaths and 78 recoveries) having all the variable data used for analysis. The design of the study is shown in Figure 1. Variables selected for the machine learning analysis were Gender, Age, Date of the onsets of symptoms, Date of Hospital Visit, Visit to the Wuhan province in China, From Wuhan Province of China, Death and Recovery. The accuracy of the model was measured by calculating the Area Under the ROC (AUROC) curve during validation on the test dataset. The AUROC on the test dataset was found to be 0.97. Age was the most important variable in the model for predicting the fate which was interestingly followed by the time gap between the onset of symptoms and hospitalization. The importance of the variables in terms of Mean Decrease in Accuracy and Mean Decrease in Gini are graphically shown in Figure 2 (A, B).

**Figure 1.**
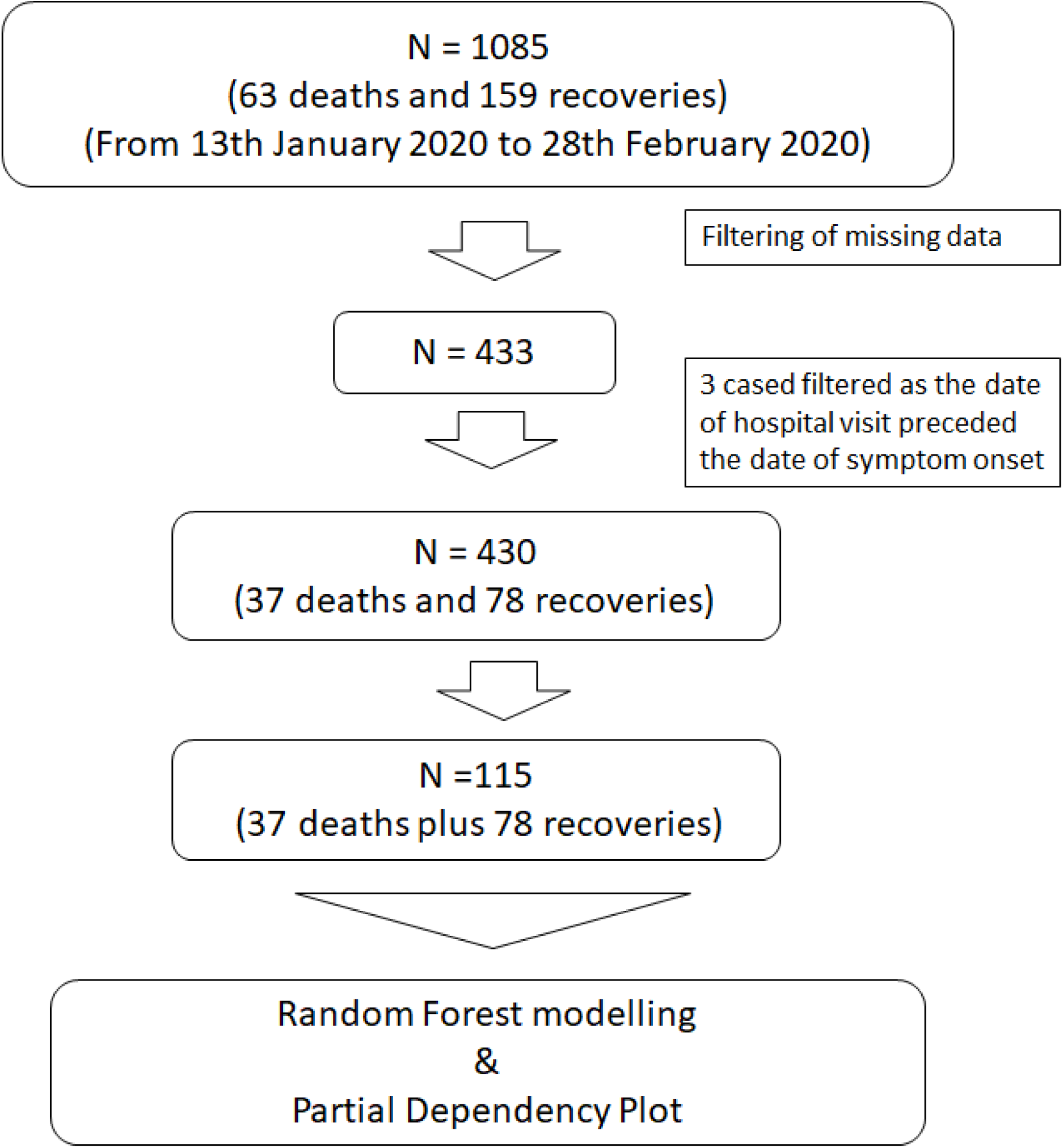
Study design. Flow diagram of Research Study.

**Figure 2.**
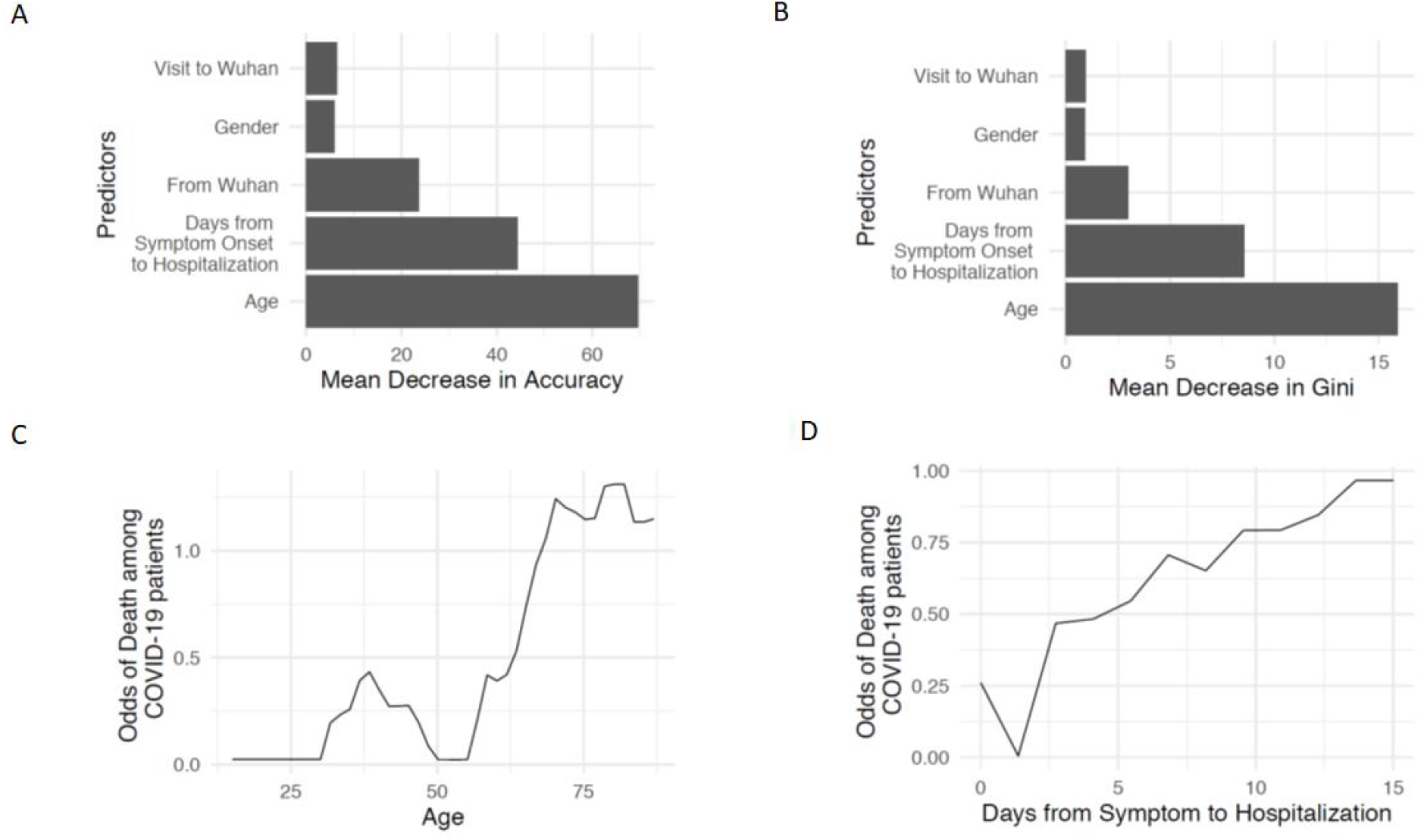
Variable Importance Plot and Partial Dependency Plot from the Random Forest model. (A, B) Variable Importance Plot showing the contribution of the predictors on the Random Forest model according to Mean Decrease in Accuracy (A) and Mean Decrease in Gini (B), Partial Dependency Plot showing the marginal effect of Age (C) and Days from Symptom to Hospitalization (D) on the odds of Death among COVID-19 patients.

In order to inspect the marginal effect of the predictors over the mortality of patients with COVID-19, we generated the partial dependency plots for the odds of Death among COVID-19 patients with Age and Days from the onset of symptoms to hospitalisation. As shown in Figure 2 (C, D), accentuation in odds of death was found with age beyond 62 years as well as beyond a time gap of 2 days between the onset of symptoms to hospitalisation. Taken together, our analysis identifies older age (>62 years) and delayed hospitalisation as the two most important predictors of mortality among patients with COVID-19.

Mortality of critically ill patients of COVID-19 is high and co-morbidities including hypertension, diabetes and coronary artery disease are often present in hospitalised patients. Though 48% of the non-survivors had a co-morbid disease, in multivariate analyses, independent associations of in-hospital death were found to be present with older age, high Sequential Organ Failure Assessment (SOFA) score and elevated d-dimer levels (6). Another study has also identified older patients as a high risk group for mortality (7). In agreement with previously published studies, our analysis also identified Age to be the most important risk factor for mortality among COVID-19 patients. However, the role of delayed hospitalisation following the development of symptoms as another significant risk factor for mortality among COVID-19 patients (after Age) is being reported for the first time. The inadequacy of healthcare resources has already been reported to associate with increased mortality among COVID-19 patients (8). However, denial, neglect and inhibition of seeking healthcare support on behalf of the patients, public shame etc. could enhance the risk of mortality. Hence, public awareness of getting healthcare service and hospitalisation at the very onset of the occurrence of symptoms could potentially reduce mortality in patients of COVID-19. Validation of these predictors of mortality in future prospective studies could be useful in identifying and stratifying the risk groups as well as provide necessary healthcare services.

## Data Availability

Raw data are available with open source Kaggle.

https://www.kaggle.com/sudalairajkumar/novel-corona-virus-2019-dataset#COVID19_line_list_data.csv

## Author Contributions

JS and PC have contributed to the concept and design of the study, data analysis, and writing of the manuscript. Authors have full access to all the data in the study and take responsibility for the integrity and the accuracy of the data analysis. Both authors have approved the final version of the article.

## Acknowledgements

JS received a research fellowship from Indian Council of Medical Research (No.3/1/3/JRF-2017/HRD-LS/56429/54).

## Disclosure Summary

The authors declare no conflict of interest.

## Notes

### Competing Interest Statement

The authors have declared no competing interest.

## REFERENCES

1. World Health Organization. Coronavirus disease (COVID-19) pandemic. https://www.who.int/emergencies/diseases/novel-coronavirus-2019 Date accessed: March 25, 2020

2. Zhou P, Yang XL, Wang XG, et al. A pneumonia outbreak associated with a new coronavirus of probable bat origin. Nature. 2020;579(7798):270–273. doi:10.1038/s41586-020-2012-7

3. Yang J, Zheng Y, Gou X, et al. Prevalence of comorbidities in the novel Wuhan coronavirus (COVID-19) infection: a systematic review and meta-analysis [published online ahead of print, 2020 Mar 12]. Int J Infect Dis. 2020;S1201-9712(20)30136-3. doi:10.1016/j.ijid.2020.03.017

4. Breiman L. Random Forests. Mach Learn 2001;45:5–32. doi:10.1023/A:1010933404324.

5. RStudio Team (2015). RStudio: Integrated Development for R. RStudio, Inc., Boston, MA url: https://www.rstudio.com

6. Zhou F, Yu T, Du R, et al. Clinical course and risk factors for mortality of adult inpatients with COVID-19 in Wuhan, China: a retrospective cohort study [published online ahead of print, 2020 Mar 11] [published correction appears in Lancet. 2020 Mar 12;:]. Lancet. 2020;S0140-6736(20)30566-3. doi:10.1016/S0140-6736(20)30566-3

7. Yang X, Yu Y, Xu J, et al. Clinical course and outcomes of critically ill patients with SARS-CoV-2 pneumonia in Wuhan, China: a single-centered, retrospective, observational study [published online ahead of print, 2020 Feb 24] [published correction appears in Lancet Respir Med. 2020 Feb 28;:]. Lancet Respir Med. 2020;S2213-2600(20)30079-5. doi:10.1016/S2213-2600(20)30079-5

8. Ji Y, Ma Z, Peppelenbosch MP, Pan Q. Potential association between COVID-19 mortality and health-care resource availability. Lancet Glob Health. 2020;8(4):e480. doi:10.1016/S2214-109X(20)30068-1

